# Prevalence evolution of SARS-CoV-2 infection in the Municipality of São Paulo, 2020 - 2021

**DOI:** 10.1101/2021.06.16.21256530

**Authors:** Jose O M Albuquerque, Gabriela A Kamioka, Geraldine Madalosso, Selma A Costa, Paula B Ferreira, Francisco A Pino, Ana Paula S Sato, Ana Carolina A Carvalho, Ana Beatriz P Amorim, Caroline C Aires, Ana Paula A G Kataoka, Elisa S M M Savani, Thirsa A F Bessa, Breno S Aguiar, Marcelo A Failla, Edson A Santos, Edjane M T Brito, Maria C H Santos, Solange M S Silva, Luiz A V Caldeira, Luiz C Zamarco, Sandra M S Fonseca, Marcia M C Lima, Ivanilda A Marques, Fabiana E V Silva, Paula R Glasser, Patrícia C P R Burihan, Cinthya L Cavazzana, Renata C Lara, Debora S Mello, Alessandra C G Pellini, Fernando Y Nishio, Fernanda M Kian, Elza S Braga, Nilza M P Bertelli, Wagner Fracini, Marcelo D A Gonçalves, Paulete S Zular, Regiane S Piva, Eduardo de Masi

## Abstract

**Objectives:** To estimate the evolution of the prevalence of SARS-CoV-2 virus infection among residents aged 18 years or over in the municipality of São Paulo.

**Methods:** This is a population-based household survey conducted every 15 days, between June and September 2020 and January and February 2021. In total, 11 phases were performed. The presence of antibodies against SARS-CoV-2 was identified in venous blood using a lateral flow test, Wondfo Biotech. In the last phase, it was combined with an immunoenzymatic test, Euroimmun. Participants also answered a semi-structured questionnaire on sociodemographic and economic factors and social distancing measures. Prevalence estimates and 95% confidence intervals were estimated according to the region, Human Development Index, sex, age group, ethnicity, education, income and variables associated with risk or prevention of the infection. To compare the frequencies among the categories of each variable, the chi-square test with Rao Scott correction was used, considering a 5% significance level.

**Results:** In total, 23,397 individuals were interviewed and had their samples collected. The estimated prevalence of antibodies against SARS-CoV-2 ranged from 9.7% (95%CI: 7.9-11.8%) to 25.0% (95%CI: 21.7-28.7). The prevalence of individuals with antibodies against SARS-CoV-2 was higher among black and pardo people, people with lower schooling, people with lower income and among residents of regions with lower Human Development Index. The lowest prevalences were associated with recommended measures of disease protection. The proportion of asymptomatic infection was 45.1%.

**Conclusion:** The estimated prevalence of SARS-CoV-2 infection was lower than the cumulative incidence variation, except for the last phase of the study. The differences in prevalence estimates observed among subpopulations showed the social inequality as a risk of infection. The lower prevalence observed among those who could follow prevention measures reinforce the need to maintain the social distancing measures as ways to prevent SARS-CoV-2 infection.

## Introduction

In December 2019, the World Health Organization (WHO) received a notification of pneumonia outbreak in Wuhan, Hubei Province, People’s Republic of China. The identification of the etiological agent occurred quickly, a new coronavirus called SARS-CoV-2. On January 30, 2020, WHO declared the disease outbreak caused by the SARS-CoV-2 virus – COVID-19 – as a Public Health Emergency of International Concern (PHEIC), the highest WHO alert level according to the International Health Regulations (IHR). On March 11, 2020, WHO declared the outbreak of COVID-19 as a global pandemic^1^.

In Brazil, the Ministry of Health (MoH) declared the COVID-19 as a Public Health Emergency of National Concern (PHENC) on February 03, 2020^2^. The first case was diagnosed on February 26, 2020 and on March 20, 2020, MoH declared the community transmission of COVID-19 in the national territory^3^.

From the date of the first case until March 28, 2021, the disease was detected in 12,490,362 people and caused 310,550 deaths, the incidence rate was 5,943.6 per 100,000 inhabitants and the mortality rate was 147.8 per 100,000 inhabitants^4^.

In the Municipality of São Paulo (MSP), until March 26, 2021, 2,390,256 cases of acute respiratory infection (ARI) in residents were notified in the report system (e-SUS Notifica), of which 609,380 (25.5%) were confirmed for SARS-CoV-2 infection. In the Epidemiological Surveillance Information System of ARI (SIVEP Gripe), 161,758 cases of severe acute respiratory syndrome (SARS), residing in the MSP were reported, of which 90,049 (55.7%) were confirmed for COVID-19. Among the COVID-19 cases, 21,051 (3%) evolved to death.

Given the epidemiological situation of COVID-19 in the MSP, on March 23, 2020 the city council adopted strategies to diminish the disease transmission, establishing voluntary quarantine with the closure of non-essential services^5^. On May 29, 2020, the Decree No. 59,473, gradually reopened some non-essential services^6^, therefore, it became extremely important to know the serological situation of the population regarding COVID-19 infection to support decision-making.

This serial serological survey was designed to represent people with 18 years or older in the MSP. Population-based data was needed to direct strategies to combat the pandemic and evaluate the effects of COVID-19 actions to prevent and control the disease. In this sense, our study aimed to estimate the prevalence of SARS-CoV-2 virus infection in adults with 18 years or older, living in the MSP; estimate the proportion of asymptomatic individuals with positive tests and describe the evolution of the prevalence of SARS-CoV-2 virus infection over time.

## Methods

This is a serial serological survey to estimate the prevalence of SARS-CoV-2 infection in the MSP.

### Study area

The MSP, the capital of the state of São Paulo, has a Human Development Index (HDI) of 0.805 and an estimated population in 2020 of 11.9 million inhabitants, of which 9.2 million were 18 years or older. The municipality presents a large economic and social disparity, reflecting heterogeneities in the education, income and housing. The Gini Index in 2010 was 0.6453^7^.

For the planning of healthcare actions, the municipality is divided into six regions: North, Central, West, Southeast, East and South, 27 Technical Health Supervisions, 472 primary healthcare units (PHU) and their respective coverage areas (CA) (Figure 1).

**Figure 1.**
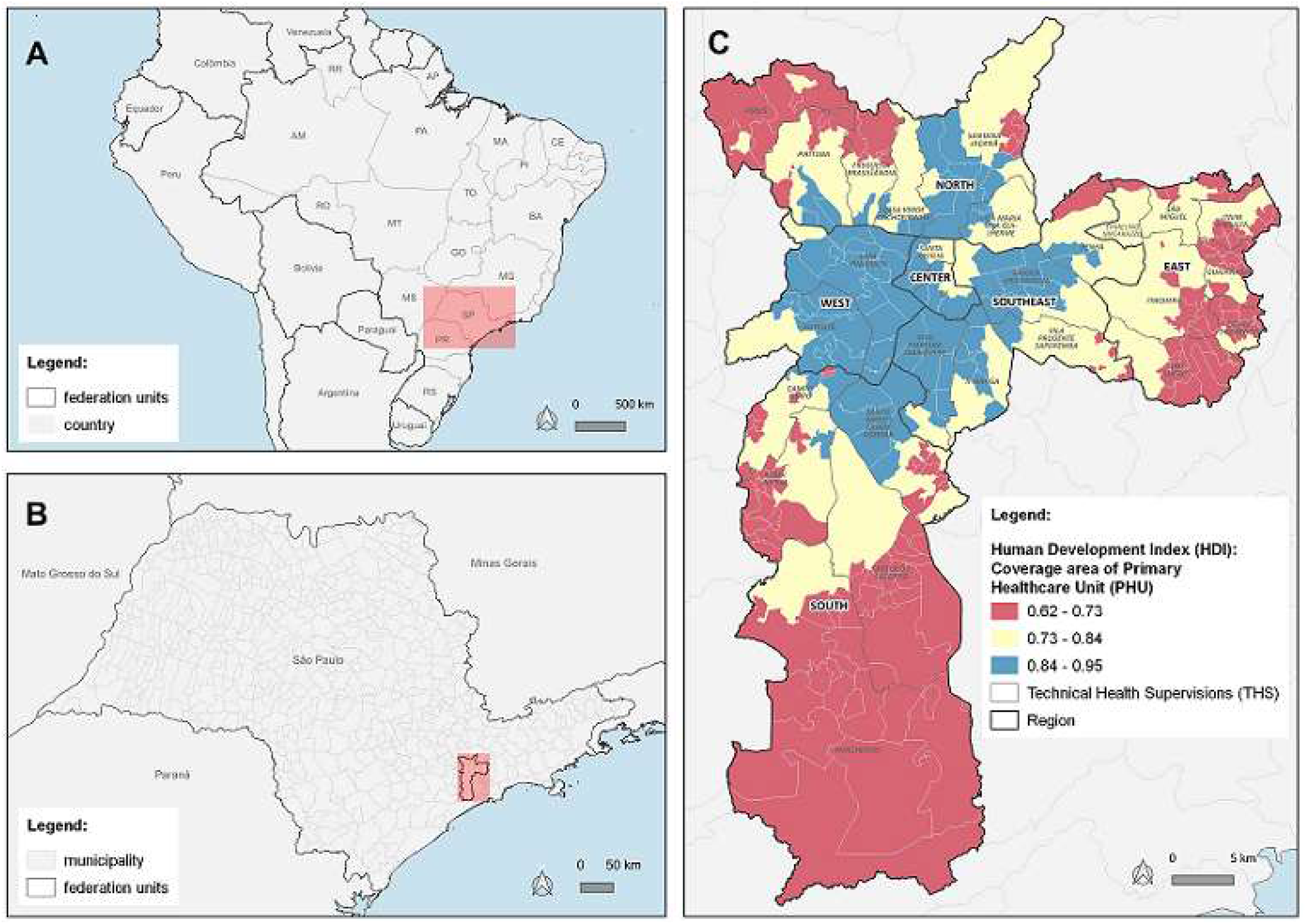
Distribution of Human Development Index (HDI) by Coverage Area of Primary Healthcare Unit (CA-PHU) and region. Municipality of São Paulo, 2020.

Figure 1 shows the social and economic disparities between and within the regions of the MSP.

### Sample methodology

In 2020, a pilot study was performed in June and seven cross-sectional studies were repeated periodically in different samples, every 15 days, from June to September. In 2021, four other studies were repeated in January and February. In each phase, a stratified probabilistic sample with a simple casual sampling was used within each stratum^8^.

For a prevalence ranging from 5% to 20%, the sample size was determined so that, the estimates were produced with coefficients of variation lower than 15% for the MSP and lower than 30% for the regions, with the exception of the Central and West regions, which have a smaller number of PHU. This process results in eight dwellings per stratum. The sample size was increased to 12 dwellings to maintain the accuracy of the estimates affected by the non-response (closed households or refusals to participate in the study). In the Central and West regions, the sample size per CA-PHU was also increased to 15 to also compensate the smaller number of PHU in the region (Table S1, supplementary material).

Statistically, this is a single stage sampling (the dwelling is the sample unit), however, operationally, the selection was made in two stages; in the second stage, the resident was selected in the household using the last birthday method. The selected person collected material for laboratory analyses and was interviewed. If the selected property was closed or the selected resident was not at home at the time of the visit, the PHU’s team should return up to twice.

### Database

The addresses of dwellings were accessed from a database composed by three different types of records updated between the study phases: a) residential property taxpayers (IPTU) of 2020; b) hydrometers of sanitation company (SABESP) of 2017; c) Family Health Strategy (FHS). The distribution of dwellings by record was unequal in the municipality. Some areas did not have representativeness from one of the records or the number was insufficient in the sample. The number of dwellings in each stratum was selected from the database, proportionally to the number of dwellings in each record.

### Testing and questionnaire

Testing was performed using the SARS-CoV-2 Antibody test® (Wondfo Biotech, Guangzhou, China) that detects IgM/IgG antibodies against SARS-CoV-2, without discriminating the type of immunoglobulin. In Brazil, the test is distributed by MoH under the name One Step COVID-2019 Test® and the legal manufacturer is Celer Biotecnologia S/A^9^.

The test is based on the principle of lateral flow immunochromatographic for the detection of IgG/IgM antibodies against SARS-CoV-2 in human blood, serum or plasma^10^.

In this survey, venous blood samples were used to obtain the serum, since the validation study of the OneStep Wondfo Test indicated an increase of sensitivity with serum sample in comparison with capillary blood obtained by digital puncture. Pellanda et al^11^ estimated a sensitivity of 84.8% and a specificity of 99.0% by assessing the results of four validated studies.

In the last phase, to increase the sensitivity and the specificity of virus detection, the samples were also processed using an immunoenzymatic test (anti-SARS-CoV-2 ELISA) from EUROIMMUN, which uses the S1 spike protein as an antigen for the detection of IgG antibodies against SARS-CoV-2 in serum^12,13^.

The official laboratory results were reported to the study participants. Participants were interviewed with a semi-structured questionnaire. The information collected were: sex, age, schooling, ethnicity (self-reported), family income, household size, symptoms potentially related to COVID-19, healthcare service use, previous SARS-CoV-2 test, contact history with suspected or confirmed cases of COVID-19. Also, the interviewees were questioned about their work regime, the social distancing adopted, facemask use, visit to non-essential places and public transportation use.

### Data analysis

Data were included in a standardized electronic form in FormSUS/DATASUS platform, version 3.0^14^. Data processing and analysis were performed using the statistical packages R and STATA version 13.

Indeterminate results were classified in the analyses as negative.

Data were analyzed considering five regions in the MSP: Central-West, East, North, Southeast and South. Prevalence estimates were weighted according to the sampling design.

Prevalence with 95% confidence intervals (CI) were estimated according to the region and HDI of the MSP, sex, age, ethnicity, education, income and presence of symptoms of the individual, risk factors, recommended measures for prevention and control of the disease and social distancing. Also, the proportion of asymptomatic infections were calculated.

For statistical data analyses, four phases were randomly chosen from the 11 phases performed in this study.

Rao Scott chi-square test was used to compare the frequencies between the categories of each variable, considering a 5% significance level.

### Ethical aspects

The study was approved by the Brazilian’s National Ethics Committee (CAAE 32947920.3.0000.0008). Blood sample were collected and the individual were interviewed only after written informed consent from all participants.

## Results

Out of 63,372 selected individuals with 18 years or over, residing in the five regions of the municipality of São Paulo (MSP), 23,397 (36.9% participation rate) were interviewed and collected samples.

Table 1 shows the prevalence estimates of antibodies against SARS-CoV-2 in the MSP and their respective 95% CI for each phase of the study. All values found were within the range of CI variation of the previous phases, except in the last phase, when two laboratory tests were used to increase the sensitivity and the specificity of virus detection.

**Table 1.** Sample size, number of sample collections, positive results and prevalence estimates of SARS-CoV-2 infection and respective 95% confidence intervals by study phase. Municipality of São Paulo, 2021.

| Study phase | Period of sample collection | Sample size | Number of sample collections | Number of positive results (SARS-CoV-2) | Prevalence estimate (95%CI) |
| --- | --- | --- | --- | --- | --- |
| Pilot | 06/10/2020 to 06/17/2020 | 5,664 | 2,645 | 247 | 9.5 (7.9-11.4) |
| 1 | 06/29/2020 to 07/02/2020 | 5,772 | 2,481 | 261 | 9.7 (7.9-11.8) |
| 2 | 07/13/2020 to 07/16/2020 | 5,760 | 2,323 | 282 | 11.1 (9.6-12.9) |
| 3 | 07/28/2020 to 07/30/2020 | 5,760 | 2,529 | 296 | 10.9 (9.2-12.9) |
| 4 | 08/11/2020 to 08/13/2020 | 5,760 | 2,447 | 298 | 11.0 (9.0-13.3) |
| 5 | 08/25/2020 to 08/27/2020 | 5,760 | 2,225 | 303 | 13.9 (11.8-16.2) |
| 6 | 09/08/2020 to 09/10/2020 | 5,760 | 2,125 | 270 | 11.9 (10.0-14.1) |
| 7 | 09/22/2020 to 09/24/2020 | 5,760 | 2,012 | 244 | 13.6 (11.6-15.8) |
| 8 | 01/05/2021 to 01/07/2021 | 5,760 | 1,908 | 278 | 14.1 (11.7-16.8) |
| 9 | 01/19/2021 to 01/21/2021 | 5,760 | 1,795 | 270 | 13.9 (11.7-16.4) |
| 10 | 02/02/2021 to 02/04/2021 | 5,760 | 1,742 | 281 | 16.0 (13.1-19.3) |
| 11 | 02/16/2021 to 02/18/2021 | 5,760 | 1,780 | 438 | 25.0 (21.7-28.7) |

Table 2 shows the prevalence estimates of SARS-CoV-2 infection in phases 1, 4, 7 and 11. The results of phase 11 with only the rapid test (11a) and with the addition of the ELISA test (11b) are presented separately.

**Table 2.** Prevalence estimates of SARS-CoV-2 infection by demographic and clinical characteristics and study phases (1, 4, 7 and 11). Municipality of São Paulo, 2021.

| Variables | Phase 1 |  |  | Phase 4 |  |  | Phase 7 |  |  | Phase 11 (a*) |  |  | Phase 11 (b*) |  |  |
| --- | --- | --- | --- | --- | --- | --- | --- | --- | --- | --- | --- | --- | --- | --- | --- |
|  | N (%) | Prevalence (95%CI) | p | N (%) | Prevalence (95%CI) | p | N (%) | Prevalence (95%CI) | p | N (%) | Prevalence (95%CI) | p | N (%) | Prevalence (95%CI) | p |
| **Region** |  |  | 0.892 |  |  | 0.027 |  |  | <0.001 |  |  | 0.027 |  |  | 0.487 |
| Central-West | 225 (14.2) | 10.1 (3.7-24.8) |  | 225 (6.3) | 5.2 (2.7-9.6) |  | 167 (4.8) | 5.5 (2.3-12.5) |  | 140 (7.8) | 9.5 (6.0-14.7) |  | 140 (9.2) | 19.4 (7.9-40.3) |  |
| East | 697 (18.6) | 10.0 (7.9-12.5) |  | 636 (19.3) | 12.3 (9.5-15.8) |  | 568 (16.4) | 11.7 (8.4-16.1) |  | 472 (24.1) | 17.8 (13.9-22.5) |  | 472 (20.5) | 26.0 (21.1-31.6) |  |
| North | 430 (14.4) | 8.5 (5.7-12.5) |  | 463 (12.3) | 8.3 (6.0-11.5) |  | 372 (16.7) | 13.8 (9.7-19.1) |  | 327 (21.6) | 19.0 (14.4-24.7) |  | 327 (17.2) | 26.0 (20.7-32.1) |  |
| Southeast | 564 (18.5) | 8.4 (5.8-12.1) |  | 501 (18.9) | 10.6 (7.5-14.8) |  | 399 (15.9) | 10.3 (6.7-15.4) |  | 363 (12.8) | 9.4 (6.0-14.2) |  | 363 (16.7) | 20.1 (16.2-26.6) |  |
| South | 565 (34.3) | 10.7 (7.8-14.6) |  | 623 (43.2) | 14.1 (9.5-20.4) |  | 510 (46.3) | 19.8 (15.7-24.8) |  | 491 (33.7) | 15.4 (10.6-22.0) |  | 491 (36.4) | 28.6 (21.7-36.7) |  |
| **Demographic** |  |  |  |  |  |  |  |  |  |  |  |  |  |  |  |
| **Age** |  |  | 0.145 |  |  | 0.241 |  |  | 0.011 |  |  | 0.347 |  |  | 0.174 |
| 18 to 34 years | 622 (28.8) | 10.0 (7.3-13.5) |  | 672 (39.6) | 13.1 (9.5-17.9) |  | 540 (21.7) | 10.8 (7.3-15.7) |  | 411 (31.1) | 16.8 (12.6-22.2) |  | 411 (31.6) | 29.4 (23.9-35.6) |  |
| 35 to 49 years | 753 (32.4) | 9.6 (5.9-15.3) |  | 680 (28.7) | 12.0 (8.1-17.6) |  | 547 (35.4) | 19.1 (14.8-24.4) |  | 489 (23.5) | 13.7 (9.0-20.3) |  | 489 (21.3) | 21.3 (16.0-27.7) |  |
| 50 to 64 years | 617 (30.0) | 12.4 (8.9-17.1) |  | 597 (17.8) | 8.1 (5.5-11.6) |  | 518 (29.5) | 14.4 (11.0-18.5) |  | 491 (21.8) | 11.6 (8.2-16.2) |  | 491 (29.3) | 27.0 (19.5-36.0) |  |
| 65 or more years | 489 (8.8) | 5.1 (2.7-9.5) |  | 499 (13.9) | 9.3 (6.1-14.0) |  | 441 (13.4) | 9.2 (5.9-14.3) |  | 402 (23.7) | 16.6 (12.8-21.2) |  | 402 (17.8) | 21.4 (17.0-26.5) |  |
| **Sex** |  |  | 0.397 |  |  | 0.264 |  |  | 0.973 |  |  | 0.053 |  |  | 0.034 |
| Men | 892 (33.8) | 8.6 (6.6-11.1) |  | 881 (33.8) | 9.4 (6.5-13.4) |  | 703 (39.2) | 13.5 (10.4-17.4) |  | 1184 (28.7) | 11.9 (9.3-15.0) |  | 1184 (29.7) | 20.7 (17.1-24.8) |  |
| Women | 1583 (66.2) | 10.1 (7.6-13.3) |  | 1567 (66.2) | 12.0 (9.5-15.1) |  | 1313 (60.8) | 13.6 (11.2-16.5) |  | 609 (71.3) | 16.0 (13.1-19.5) |  | 609 (70.3) | 27.3 (22.7-32.6) |  |
| Unknown | 6 (-) | - |  | 0 (-) | - |  | 0 (-) | - |  | 0 (-) | - |  | 0 (-) | - |  |
| **Race/color** |  |  | 0.029 |  |  | 0.002 |  |  | <0.001 |  |  | 0.005 |  |  | 0.018 |
| White | 1292 (40.7) | 7.3 (5.5-9.5) |  | 1263 (36.3) | 7.5 (5.7-9.9) |  | 1002 (37.4) | 9.5 (7.2-12.4) |  | 827 (39.9) | 11.6 (9.5-14.1) |  | 827 (43.2) | 21.6 (16.7-27.4) |  |
| Black/Pardo | 1137 (58.0) | 12.7 (9.5-16.9) |  | 1128 (62.0) | 15.1 (11.6-19.6) |  | 969 (61.4) | 18.8 (15.5-22.7) |  | 924 (59.7) | 18.0 (14.2-22.6) |  | 924 (56.4) | 29.3 (24.8-34.2) |  |
| Asian | 42 (1.3) | 7.1 (1.2-33.1) |  | 41 (1.8) | 13.7 (2.9-46.1) |  | 23 (1.3) | 15.1 (4.2-41.8) |  | 23 (0.4) | 3.8 (0.0-29.4) |  | 23 (0.4) | 6.2 (1.1-28.0) |  |
| Indigenous | 4 (0.0) | - |  | 4 (0.0) | - |  | 1 (0.0) | - |  | 1 (0.0) | - |  | 1 (0.0) | - |  |
| Unknown | 6 (-) | - |  | 12 (-) | - |  | 21(-) | - |  | 18 (-) | - |  | 18 (-) | - |  |
| **Education level** |  |  | <0.001 |  |  | 0.003 |  |  | <0.001 |  |  | 0.186 |  |  | 0.094 |
| Middle School | 764 (39.7) | 14.9 (10.4-20.8) |  | 730 (33.6) | 15.6 (11.4-21.1) |  | 661 (37.2) | 17.8 (14.3-22.0) |  | 565 (32.1) | 15.9 (10.7-23.0) |  | 565 (34.4) | 29.1 (21.0-38.9) |  |
| High school | 1007 (35.1) | 8.6 (6.7-10.8) |  | 963 (46.8) | 11.9 (8.6-16.2) |  | 803 (48.0) | 16.0 (12.5-20.1) |  | 684 (40.7) | 15.7 (12.4-19.7) |  | 684 (39.3) | 26.0 (21.7-30.7) |  |
| Higher education | 609 (16.3) | 4.9 (3.1-7.4) |  | 580 (16.1) | 5.9 (3.9-8.9) |  | 409 (11.5) | 6.0 (3.6-9.7) |  | 351 (20.3) | 11.2 (8.1-15.1) |  | 351 (20.6) | 19.4 (15.2-24.4) |  |
| Illiterate | 87 (9.0) | 29.5 (11.6-57.0) |  | 122 (3.6) | 9.4 (4.1-19.7) |  | 78 (3.3) | 14.7 (5.7-33.0) |  | 94 (7.0) | 24.7 (12.8-42.4) |  | 94 (5.8) | 35.5 (21.8-52.1) |  |
| Unknown | 14 (-) | - |  | 53 (-) | - |  | 65 (-) | - |  | 99 (-) | - |  | 99 (-) | - |  |
| **Clinics** |  |  |  |  |  |  |  |  |  |  |  |  |  |  |  |
| Asymptomatic |  |  | <0.001 |  |  | <0.001 |  |  | <0.001 |  |  | <0.001 |  |  | <0.001 |
| Yes | 1708 (43.6) | 6.0 (4.0-8.8) |  | 1609 (40.5) | 6.4 (4.5-9.1) |  | 1351 (35.3) | 7.5 (5.6-10.1) |  | 1111 (45.7) | 11.3 (8.3-15.1) |  | 1111 (45.1) | 19.0 (14.9-23.8) |  |
| No | 766 (56.4) | 18.3 (14.7-22.6) |  | 838 (59.5) | 20.7 (16.4-25.8) |  | 661 (64.7) | 24.6 (20.7-28.9) |  | 601 (54.3) | 21.1 (17.4-25.5) |  | 601 (54.9) | 36.4 (30.6-42.6) |  |
| Unknown | 7 (-) | - |  | 1 (-) | - |  | 4 (-) | - |  | 81 (-) | - |  | 81 (-) | - |  |

(a)\* = rapid test; (b)\* = rapid test + ELISA.

The prevalence estimates of this study did not follow the evolution of the accumulated cases of SARS-CoV-2 infection in the MSP, except for the last phase as shown in Figure 2A.

**Figure 2.**
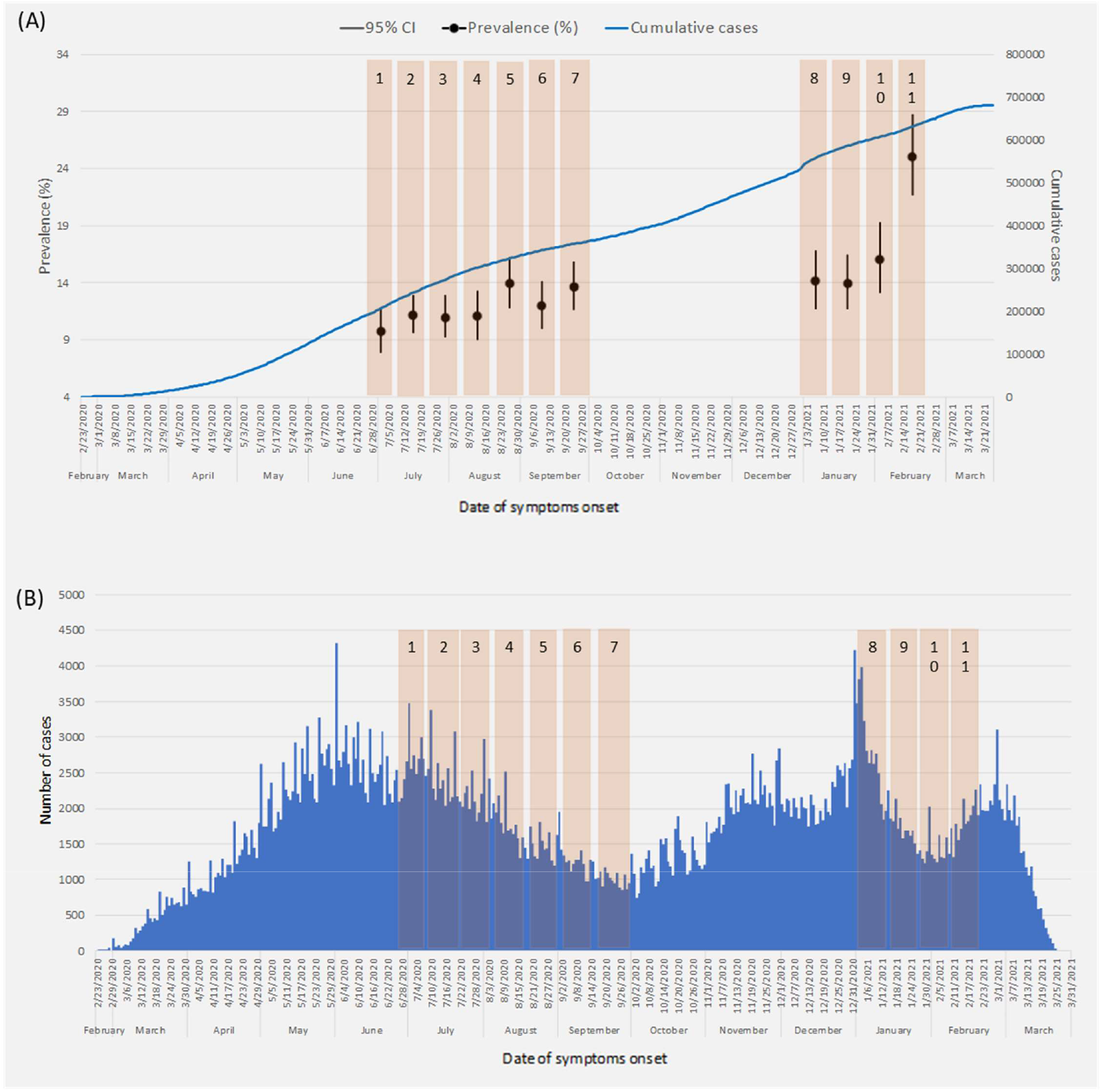
(A) Prevalence estimates and 95% confidence interval of SARS-CoV-2 infection by study phases; cumulative cases of SARS-CoV-2 infection by date of symptoms onset; (B) Distribution of COVID-19 cases by date of symptoms onset and study phases. Municipality of São Paulo, 2021.

The design effect (deff) varied between the study phases, from 1.93 in phase 7 to 3.09 in the last phase; in phase 1, the deff was 2.71 and in phase 4; 2.95.

The coefficients of variation of prevalence estimates for the municipality were below 15% in all phases and below 30% in all regions, except for the Central-West region.

There was a significant difference between the regions of the municipality of São Paulo in phases 4 and 7 of the study. The Central-West region presented the lowest estimates, whereas the Southern region presented the highest.

Table 2 also presents the prevalence estimates of SARS-CoV-2 infection, according to demographic and clinical characteristics. Regarding age, the estimated prevalences varied between the phases of the study, in phase 1, the highest prevalence was found in the age group from 50 to 64 years; in phase 4, from 18 to 34 years; in phase 7, from 35 to 49 years and in phase 11, from 18 to 34 years. Only in phase 7, the difference among age groups was significant.

In all phases, women presented the highest prevalence, especially in phase 11, when the difference was statistically significant. The estimated prevalence of black and pardo people was the highest in all phases. There was a significant difference in ethnicity in all phases.

The highest prevalence estimates were found in the groups with the lowest education levels. Table 2 shows significant differences between the education levels in phases 1, 4 and 7.

The proportion of asymptomatic individuals among the positive cases ranged from 35.3% (95%CI: 29.5-41.6) in phase 7 to 45.1% (95%CI: 38.2-52.2) in phase 11. In phase 1, the proportion was 43.6% (95%CI: 37.4-50.0) and in phase 4, 40.5% (95%CI: 31.8-49.9). As shown in Table 2, the prevalence of cases without symptoms increased throughout the study phases.

Table 3 shows the prevalence estimates of SARS-CoV-2 infection, according to socioeconomic factors and recommended measures adopted, for phases 1, 4, 7 and 11.

**Table 3.** Prevalence estimates of SARS-CoV-2 infection by socioeconomic factors. recommended measures and study phases (1. 4. 7 and 11). Municipality of São Paulo, 2021.

| Variables | Phase 1 |  |  | Phase 4 |  |  | Phase 7 |  |  | Phase 11 (a*) |  |  | Phase 11 (b*) |  |  |
| --- | --- | --- | --- | --- | --- | --- | --- | --- | --- | --- | --- | --- | --- | --- | --- |
|  | N (%) | Prevalence (95%CI) | p | N (%) | Prevalence (95%CI) | p | N (%) | Prevalence (95%CI) | p | N (%) | Prevalence (95%CI) | p | N (%) | Prevalence (95%CI) | p |
| **Economic** |  |  |  |  |  |  |  |  |  |  |  |  |  |  |  |
| **HDI** |  |  | 0.015 |  |  | 0.021 |  |  | <0.001 |  |  | 0.068 |  |  | 0.281 |
| Range A (0.84 to 0.95) | 390 (22.6) | 8.3 (4.2-15.7) |  | 347 (14.8) | 6.2 (3.8-10.0) |  | 270 (8.7) | 4.6 (2.7-8.0) |  | 204 (16.8) | 10.7 (7.1-15.7) |  | 204 (18.5) | 20.1 (12.3-30.9) |  |
| Range B (0.73 to 0.84) | 1401 (39.6) | 7.5 (5.7-9.9) |  | 1304 (54.7) | 12.1 (9.3-15.6) |  | 1038 (54.8) | 15.3 (12.1-19.1) |  | 978 (50.1) | 14.4 (11.2-18.3) |  | 978 (51.5) | 25.4 (20.8-30.6) |  |
| Range C (0.62 to 0.73) | 690 (37.8) | 16.2 (12.9-20.1) |  | 797 (30.5) | 14.0 (9.7-19.8) |  | 708 (36.4) | 19.3 (15.7-23.4) |  | 611 (33.1) | 18.4 (14.3-23.4) |  | 611 (30.1) | 28.8 (24.0-34.1) |  |
| **Income** |  |  | 0.040 |  |  | <0.001 |  |  | 0.037 |  |  | 0.054 |  |  | 0.005 |
| Class A/B | 132 (2.1) | 2.3 (0.6-8.7) |  | 144 (3.3) | 4.4 (1.7-10.8) |  | 77 (1.8) | 8.9 (1.3-42.5) |  | 64 (1.8) | 3.9 (0.9-15.2) |  | 64 (1.2) | 4.7 (1.3-15.0) |  |
| Class C | 1182 (47.5) | 8.9 (6.2-12.7) |  | 1013 (34.2) | 8.3 (6.5-10.7) |  | 842 (35.8) | 10.2 (7.9-13.1) |  | 763 (45.6) | 12.7 (9.9-16.0) |  | 763 (48.3) | 23.7 (18.8-29.4) |  |
| Class D/E | 974 (41.3) | 12.5 (9.5-16.2) |  | 1055 (57.3) | 18.2 (13.7-23.7) |  | 913 (52.9) | 18.9 (15.5-23.0) |  | 757 (42.9) | 17.9 (13.3-26.7) |  | 757 (40.5) | 29.8 (23.7-36.7) |  |
| Not informed | 193 (9.2) | 10.3 (6.1-16.8) |  | 211 (5.2) | 4.7 (2.4-9.1) |  | 160 (9.5) | 11.4 (5.5-21.8) |  | 142 (9.7) | 13.0 (0.7-22.6) |  | 142 (10.0) | 23.4 (15.7-33.4) |  |
| Unknown | 21 (-) | - |  | 25 (-) | - |  | 24 (-) | - |  | 67 (-) | - |  | 67 (-) | - |  |
| **Recommended measures** |  |  |  |  |  |  |  |  |  |  |  |  |  |  |  |
| **Work situation** |  |  | <0.001 |  |  | 0.002 |  |  | 0.097 |  |  | 0.593 |  |  | 0.121 |
| Unemployed | 411 (26.6) | 15.4 (9.0-25.0) |  | 427 (29.8) | 18.1 (11.6-27.1) |  | 340 (15.3) | 13.8 (9.2-20.0) |  | 291 (15.5) | 14.7 (9.8-21.3) |  | 291 (14.4) | 23.4 (17.1-31.3) |  |
| Telework | 439 (8.5) | 3.9 (2.6-6.0) |  | 381 (8.0) | 4.4 (2.2-8.7) |  | 277 (10.5) | 8.7 (4.8-15.0) |  | 213 (13.5) | 13.6 (8.5-21.1) |  | 213 (11.4) | 19.8 (14.1-27.2) |  |
| Employed | 591 (33.6) | 14.1 (10.7-18.3) |  | 613 (26.6) | 11.9 (9.1-15.5) |  | 535 (33.5) | 18.1 (13.7-23.5) |  | 519 (35.3) | 17.2 (12.5-23.2) |  | 519 (37.0) | 31.0 (24.3-38.7) |  |
| Not active | 874 (25.1) | 7.4 (4.9-11.1) |  | 909 (31.8) | 10.6 (7.5-14.7) |  | 770 (37.6) | 13.5 (10.9-16.5) |  | 665 (32.7) | 13.6 (10.5-17.4) |  | 665 (33.8) | 24.1 (18.7-30.5) |  |
| Mixed work | 134 (6.2) | 10.3 (6.0-17.2) |  | 107 (3.9) | 9.0 (4.5-17.2) |  | 81 (3.1) | 7.7 (2.2-23.4) |  | 72 (3.0) | 9.8 (4.8-19.0) |  | 72 (3.4) | 19.6 (11.7-30.9) |  |
| Unknown | 32 (-) | - |  | 11 (-) | - |  | 13 (-) | - |  | 33 (-) | - |  | 33 (-) | - |  |
| **Use of public transport** |  |  | 0.013 |  |  | 0.607 |  |  | 0.437 |  |  | 0.249 |  |  | 0.022 |
| No | 1944 (75.0) | 9.2 (7.0-11.8) |  | 1755 (74.0) | 11.3 (8.9-14.1) |  | 1367 (66.7) | 13.6 (11.4-16.2) |  | 969 (53.5) | 12.8 (10.3-15.7) |  | 969 (50.7) | 20.2 (17.2-23.6) |  |
| Yes | 446 (25.0) | 14.0 (10.9-17.7) |  | 586 (26.0) | 10.3 (7.7-13.6) |  | 575 (33.3) | 15.5 (11.7-20.4) |  | 661 (46.5) | 15.8 (11.6-21.2) |  | 661 (49.3) | 28.0 (22.1-34.7) |  |
| Unknown | 91 (-) | - |  | 107(-) | - |  | 74 (-) | - |  | 163 (-) | - |  | 163 (-) | - |  |
| **Social distancing** |  |  | <0.005 |  |  | 0.342 |  |  | 0.320 |  |  | 0.006 |  |  | 0.005 |
| Totally | 1700 (58.4) | 8.0 (5.9-10.1) |  | 1871 (73.9) | 10.3 (8.1-12.9) |  | 1566 (70.3) | 12.7 (10.6-15.1) |  | 1260 (66.3) | 13.0 (10.4-16.2) |  | 1260 (66.8) | 22.6 (18.4-27.4) |  |
| Partially | 701 (37.8) | 13.6 (10.7-17.1) |  | 530 (24.3) | 13.1 (8.8-19.0) |  | 412 (27.9) | 17.6 (13.2-23.1) |  | 475 (32.9) | 19.5 (15.8-23.9) |  | 475 (32.2) | 32.9 (28.1-38.1) |  |
| Not adopted | 41 (3.8) | 18.9 (6.9-42.1) |  | 35 (1.8) | 17.9 (6.3-41.2) |  | 24 (1.8) | 20.0 (2.5-70.6) |  | 22 (0.8) | 10.0 (5.4-17.7) |  | 22 (1.1) | 23.8 (8.7-50.4) |  |
| Unknown | 39 (-) | - |  | 12 (-) | - |  | 14 (-) | - |  | 36 (-) | - |  | 36 (-) | - |  |
| **Use of mask** |  |  |  |  |  | 0.677 |  |  | 0.841 |  |  | 0.155 |  |  | 0.022 |
| Always |  |  |  | 2108 (87.4) | 11.0 (8.9-13.7) |  | 1734 (89.1) | 13.6 (11.6-15.9) |  | 1496 (82.4) | 13.9 (11.7-16.6) |  | 1496 (84.1) | 24.4 (20.7-28.6) |  |
| Mostly | Data obtained from phase 2 |  |  | 225 (9.7) | 10.9 (6.6-17.6) |  | 185 (7.6) | 13.2 (6.9-23.7) |  | 178 (13.8) | 22.7 (14.8-33.1) |  | 178 (11.1) | 31.6 (22.6-42.2) |  |
| Sometimes |  |  |  | 68 (2.6) | 15.3 (6.5-31.8) |  | 53 (3.0) | 21.2 (6.1-52.7) |  | 58 (3.5) | 16.9 (6.3-38.2) |  | 58 (3.6) | 29.4 (13.8-52.0) |  |
| Never |  |  |  | 9 (0.3) | 7.5 (0.6-48.9) |  | 7 (0.3) | - |  | 5 (0.3) | 3.7 (0.9-13.9) |  | 5 (1.2) | 85.0 (37.7-98.1) |  |
| Unknown |  |  |  | 17 (-) | - |  | 27 (-) | - |  | 41 (-) | - |  | 41 (-) | - |  |

(a)\* = rapid test; (b)\* = rapid test + ELISA.

Prevalence estimates were inverse to the categories of family income that were distributed in A/B (≥ R$ 8,641.00), C (R$ 2,005.00 to R$ 8,640.00) and D/E (≤ R$ 2,004.00). Moreover, HDI ranges were used to estimate the regional differences of the city, range A (0.84 to 0.95) had the lowest prevalence estimates whereas range C (0.62 to 0.73) presented the highest estimates. The difference among HDI ranges and family income categories were significantly different, except in phase 11 (Table 3).

Regarding the work situation of the interviewees, the study showed that those in telework had lower prevalence when compared to the other categories, especially in phases 1 and 4 (3.9% and 4.4%, respectively). Likewise, those who did not use public transportation and practiced social distancing presented the lowest prevalence estimates. The difference was significantly in phases 1 and 11.

The difference between the categories for mask use, “always”, “most of the time” and “sometimes” was significant in phase 11 (24.4%).

Table S2 (supplementary material) shows the reasons for non-response in each phase of the study. In 62.6% (39,169) of the selected dwellings it was not possible to conduct the interview and sample collection. The non-response rate presented an increase trend throughout the study.

## Discussion

The results showed that most people in MSP are still susceptible to the SARS-CoV-2 virus even with the increasing number of SARS-CoV-2 infections during the pandemic. The prevalence of individuals with positive results was higher among the black and pardo people when compared with the white people. Also, the prevalence was inversely associated with the education level and the income category of the individual and with the HDI of the primary healthcare unit coverage area (CA-PHU) of the selected dwelling. The lowest prevalences were associated with recommended protection measures against coronavirus disease. In 2020, several studies in Brazil and around the world aimed to estimate the prevalence of SARS-CoV-2 infection in their populations^15-21^. The results of different prevalence studies should be carefully compared, since they depend on the moment of the pandemic evolution and the laboratory test used^22^.

In our study, the estimated prevalence of antibodies against SARS-CoV-2 in the MSP ranged from 9.7% (95%CI: 7.9-11.8) in phase 1 to 16% (95%CI: 13.1-19.3) in phase 10; in phase 11, when two laboratory tests were used, the estimate was 25% (95%CI: 21.7-28.7). In Brazil, Hallal et al.^20^ conducted a large study involving two serological surveys in 133 sentinel cities in all Brazilian states, they used the same laboratory test of our study with finger prick blood samples and estimated prevalences between 0% and 25%. In the state of Espírito Santo, Gomes et al.^22^ conducted a population-based serial cross-sectional study in 11 municipalities, they found a prevalence of 2.1% using the Celer Technologies Inc test in a finger prick blood sample. In the municipality of São Paulo, a study conducted in six administrative districts found a low prevalence (4.7%) using the MAGLUMI 2019-nCoV chemiluminescence immunoassay test^21^.

In the present study, the highest prevalence estimates were initially found in the age group from 50 to 64 years, which is a vulnerable population most affected at the beginning of the pandemic. In the following phases, the younger age groups were the most prevalent, which is the economically active population that most circulates in the city. This fact may be associated with the economic recover of the municipality with the reduction of restrictions and reopening of commercial establishments^6^.

Similar to other studies, there was no difference between the prevalence of men and women ^16,17,20^.

Disparities in prevalence estimates by ethnicity and level of education are consistent with the inequalities associated with demographic, socioeconomic and risk factors for the disease transmission in the studied municipality, as shown by Tess et al.^21^ and Rosenberg et al.^15^.

The prevalence of SARS-CoV-2 infection was inverse to the individual’s income and to the HDI of CA-PHU, therefore, the lower the income and HDI in the CA-PHU, the higher the prevalence of SARS-CoV-2 infection. Similar to the studies of Bermudi et al.^23^ and Menezes et al.^24^, vulnerable populations with lower income and poor housing conditions have a higher risk of virus transmission and difficulties to acess healthcare for diagnosis and treatment.

The proportion of individuals that have not reported symptoms related to COVID-19 since the beginning of March 2020 among those with positive test for the SARS-CoV-2 virus varied among the study phases. The proportion of up to 45.1% was high when compared with other studies. The proportion of asymptomatic individuals with 18 years or older reported in seroprevalence studies in England^18^ and in a USA city (Chelsea)^25^ were 32.2% and 24.7%, respectively. Similar to this study, Tess et al.^21^ found a high proportion of asymptomatic individuals in residents of six districts of the MSP (45.3%).

The risk associated with the work situation was lower among individuals working from home when compared to those who work at the office, this results were consistent with other seroprevalence studies of SARS-CoV-2^26,27^.

The use of masks was a protective measure against SARS-CoV-2 infection, similar to that found in studies performed in the state of Maranhão^26^ and in China^28^.

This study has some limitations. It included the high rate of non-response, the address or the dwelling selected in the database were not identified (29.0%) or the selected individual refused to receive the work team or participate in the study (26.7%), since venous blood samples were collected instead of finger prick in order to increase the sensitivity of the test, as shown by Hallal et al.^20^ and Tess et al.^21^ The large number of teams (471) composed of employees of the primary healthcare units and responsible for data collection may have contributed to divergences in the approach of individuals to undergo the interview and the sample collection. The design effect (deff) observed for the prevalence estimates for the municipality and for the regions was higher than expected for stratified samples, possibly due to the large number of strata in the study^8^. The high deff contributed to the low accuracy of the estimates; therefore, future studies should review the sample design. Individuals under 18 years of age were not included in the sample. The use of a second test in phase 11 enhanced the sensitivity of the test. ELISA test detected 53.8% more positive cases than the rapid test. Possibly, the prevalence estimates may have been underestimated in phases 1 to 10.

Serological tests may present false-negative results in the first days of the infection, therefore, it has little diagnostic value for acute cases. The proportion of negative cases among symptomatic individuals with sample collection performed up to the 14th day of the date of symptoms onset decreased throughout the study, from 18.7% in phase 1 to 10.1% in the last phase. The probability of selecting individuals during the first 14 days of infection in the sample was decreasing and the recall bias of the date of symptoms onset was 16.4%.

In conclusion, the prevalence variation of SARS-CoV-2 infection was lower than the variation of the cumulative incidence rate, except for the last phase of the study. The differences in prevalence estimates according to protective measures against SARS-CoV-2 infection reinforce the need to maintain social distancing, mask use and telework in all age groups and social classes.

Sequential phases will allow the monitoring of the pandemic evolution and will verify the effectiveness of the current recommended protection measures for the population. Further studies should consider vaccinated people and the influence of vaccination on the population.

## Data Availability

The data that support the findings of this study are available on request from the corresponding author, SMS/COVISA. The data are not publicly available due to restrictions of their containing information that could compromise the privacy of research participants.

## Notes

### Competing Interest Statement

The authors have declared no competing interest.

### Funding Statement

No external funding was received.

### Author Declarations

The project was approved by Brazilian's National Ethics Commitee (CAAE 32947920.3.0000.0008).

## References

1. World Health Organization. WHO Coronavirus disease (COVID-19) Pandemic. Geneva; 2020. Available from: https://www.who.int/emergencies/diseases/novel-coronavirus-2019

2. Ministério da Saúde (Brasil). Portaria nº 188 de 3 de fevereiro de 2020. Declara Emergência em Saúde Pública de importância Nacional (ESPIN) em decorrência da Infecção Humana pelo novo Coronavírus (2019-nCoV). Diário Oficial da União, Brasília, 04 fev. 2020. Seção 1, p.1.

3. Ministério da Saúde (Brasil). Portaria nº 454 de 20 de março de 2020. Declara, em todo o território nacional, o estado de transmissão comunitária do coronavírus (COVID-19). Diário Oficial da União, Brasília, 20 mar. 2020. Seção 1, p.1.

4. Ministério da Saúde (BR). Painel Coronavírus. Atualização diária. Brasília, DF; 2021. Available from: https://covid.saude.gov.br/

5. São Paulo (Município). Decreto Municipal nº 59.298 de 23 de março de 2020. Suspende o atendimento presencial ao público em estabelecimentos comerciais e de prestação de serviços. Diário Oficial da Cidade de São Paulo, São Paulo, 23 mar. 2020a. Seção 1, p.1.

6. São Paulo (Município). Decreto Municipal nº 59.473 de 29 de maio de 2020. Estabelece, nos termos do Decreto Estadual nº 64.994, de 28 de maio de 2020, normas para o funcionamento de estabelecimentos de comércio e de serviços localizados na Cidade de São Paulo. Diário Oficial da Cidade de São Paulo, São Paulo, 29 mai. 2020b. Seção 1, p.1.

7. DATASUS. Tecnologia da Informação a Serviço do SUS. Available from: http://www2.datasus.gov.br/DATASUS/index.php?area=02

8. Kish L. Survey Sampling. New York, Wiley, 1965.

9. Castro R, Luz P M, Wakimoto M D, Veloso V G, Grinsztejn B, Perazzo H. COVID-19: a meta-analysis of diagnostic test accuracy of commercial assays registered in Brazil. Braz J Infect Dis. 2020;968:1–8. http://dx.doi.org/10.1016/j.bjid.2020.04.003

10. Clinical General Report. SARS-CoV-2 Antibody Test (Lateral Flow Method). Guangzhou Wondfo Biotech Co., Ltd. No. 8 Lizhishan Road, Science City, Luogang District, 510663, Guangzhou, P.R. China. Available from: https://www.wondfo.com.cn

11. Pellanda LC, Wendland EMR, McBride AJA, Tovo-Rodrigues L, Ferreira MRA, Dellagostin OA et al. Sensitivity and specificity of a rapid test for assessment of exposure to SARS-CoV-2 in a community-based setting in Brazil. medRxiv [Preprint]. 2020. https://doi.org/10.1101/2020.05.06.20093476

12. Euroimmun. Coronavirus. COVID-19. Metodologia ELISA. Available from: https://brasil.euroimmun.com.br/coronavirus

13. Kubina R, Dziedzic A. Molecular and Serological Tests for COVID-19 a Comparative Review of SARS-CoV-2 Coronavirus Laboratory and Point-of-Care Diagnostics. Diagnostics (Basel). 2020;10(6):434. https://doi.org/10.3390/diagnostics10060434

14. FormSus®-Versão 3.0. DATASUS [Internet]. Available from: https://formsus.datasus.gov.br/site/default.php

15. Rosenberg ES, Tesoriero JM, Rosenthal EM, Chung R, Barranco MA, Styer LM et al. Cumulative incidence and diagnosis of SARS-CoV-2 infection in New York. Ann Epidemiol. 2020;48:23–29. https://doi.org/10.1016/j.annepidem.2020.06.00

16. Pollán M, Pérez-Gómez B, Pastor-Barriuso R, Oteo J, Hernán MA, Pérez-Olmeda M et al. on behalf of the ENE-COVID Study Group. Prevalence of SARS-CoV-2 in Spain (ENE-COVID): a nationwide, population-based sero epidemiological study. Lancet. 2020;396:535–44. https://doi.org/10.1016/S0140-6736(20)31483-5

17. Stringhini S, Wisniak A, Piumatti G, Azman AS, Lauer SA, Baysson H et al. Seroprevalence of anti-SARS-CoV-2 IgG antibodies in Geneva, Switzerland (SEROCoV-POP): a population-based study. Lancet 2020; 396:313–19. https://doi.org/10.1016/S0140-6736(20)31304-0

18. Ward H, Atchison C, Whitaker M, Ainslie KEC, Elliott J, Okell L, et al. Antibody prevalence for SARS-CoV-2 following the peak of the pandemic in England: REACT2 study in 100,000 adults. medRxiv [Preprint]. 2020. https://doi.org/10.1101/2020.08.12.20173690.

19. Liu A, Li Y, Wan Z, Wang W, Lei X, Lv Y. Seropositive Prevalence of Antibodies Against SARS-CoV-2 in Wuhan, China. JAMA Netw Open. 2020;3(10):e2025717. https://doi.org/10.1001/jamanetworkopen.2020.25717

20. Hallal P, Hartwig FP, Horta BL, Silveira MF, Struchiner CJ, Vidaletti LP, et al. SARS-CoV-2 antibody prevalence in Brazil: results from two successive nationwide serological household surveys. Lancet Glob Health. 2020;8(11): e1390–e1398. https://doi.org/10.1016/S2214-109X(20)30387-9

21. Tess BH, Granato CFH, Alves MCGP, Pintao MC, Rizzatti E, Nunes MC, Reinach FC. SARS-CoV-2 seroprevalence in the municipality of São Paulo, Brazil, ten weeks after the first reported case. medRxiv [Preprint]. 2020. https://doi.org/10.1101/2020.06.29.20142331

22. Gomes CC, Junior CC, Zandonade E, Maciel ELN, de Alencar FEC, Almada GL et al. A population-based study of the prevalence of COVID-19 infection in Espírito Santo, Brazil: methodology and results of the first stage. medRxiv [Preprint]. 2020. https://doi.org/10.1101/2020.06.13.20130559

23. Bermudi PMM, Lorenz C, Aguiar BS, Failla MA, Barrozo LV, Chiaravalloti-Neto F. Spatiotemporal ecological study of COVID-19 mortality in the city of São Paulo, Brazil: Shifting of the high mortality risk from areas with the best to those with the worst socio-economic conditions. Travel Med Infect Dis. 2021;39:101945. https://doi.org/10.1016/j.tmaid.2020.101945

24. Menezes AMB, Victora CG, Hartwig FP, Silveira MF, Horta BL, Barros AJD et al. High prevalence of symptoms among Brazilian subjects with antibodies against SARS-CoV-2: a nationwide household survey. medRxiv [Preprint]. 2020. https://doi.org/10.1101/2020.08.10.20171942

25. Naranbhai V, Chang CC, Beltran WFG, Miller TE, Astudillo MG, Villalba JA et al. High Seroprevalence of Anti-SARS-CoV-2 Antibodies in Chelsea, Massachusetts. J Infect Dis. 2020;222(12):1955–1959. https://doi.org/10.1093/infdis/jiaa579

26. Silva AAM, LimaNeto LG, Azevedo CMPS, Costa LMM, Bragança MLBM, Barros Filho AKD, et al. Population-based seroprevalence of SARS-CoV-2 and the herd immunity threshold in Maranhão. Rev Saude Publica. 2020;54:131. http://dx.doi.org/10.11606/s1518-8787.2020054003278

27. Lastrucci V, Lorini C, Del Riccio M, Gori E, Chiesi F, Sartor G et al. SARS-CoV-2 Seroprevalence Survey in People Involved in Different Essential Activities during the General Lock-Down Phase in the Province of Prato (Tuscany, Italy). Vaccines (Basel). 2020;8(4):778. http://dx.doi.org/10.3390/vaccines8040778

28. Chen Y, Tong X, Wang J, Huang W, Yin S, Huang R et al. High SARS-CoV-2 antibody prevalence among healthcare workers exposed to COVID-19 patients. J Infect. 2020;81(3):420–426. http://dx.doi.org/10.1016/j.jinf.2020.05.06

