## Supplementary material for "Prevalence evolution of SARS-CoV-2 infection in the Municipality of São Paulo, 2020 - 2021"

Table S1. Number of Primary Healthcare Units (PHU), initial and increased sample size by region. Municipality of São Paulo, 2020.

| Region | Number of PHU | Initial sample size | Increased sample size to compensate for the non-response (50%) |
|---|---|---|---|
| Central-West | 40 | 400 | 600 |
| North | 94 | 752 | 1,128 |
| South | 124 | 992 | 1,488 |
| Southeast | 95 | 760 | 1,140 |
| East | 118 | 944 | 1,416 |
| Municipality of São Paulo | 471 | 3,848 | 5,772 |

Table S2. Reasons for non-response of selected dwellings. Municipality of São Paulo, 2021.

| Study phase | Address not found | | Closed property | | Individual's refuse | | Absence of selected individual | | Vaccinated individual (COVID-19) | | Other reasons | | Total |
|---|---|---|---|---|---|---|---|---|---|---|---|---|---|
| | N | % | N | % | N | % | N | % | N | % | N | % | |
| Phase 1 | 1,234 | 39.7 | 403 | 13.0 | 774 | 24.9 | 272 | 8.8 | - | - | 422 | 13.6 | 3,105 |
| Phase 2 | 1,288 | 37.9 | 433 | 12.7 | 829 | 24.4 | 380 | 11.2 | - | - | 470 | 13.8 | 3,400 |
| Phase 3 | 1,005 | 31.7 | 533 | 16.8 | 843 | 26.6 | 425 | 13.4 | - | - | 365 | 11.5 | 3,171 |
| Phase 4 | 933 | 28.3 | 529 | 16.1 | 877 | 26.6 | 584 | 17.7 | - | - | 371 | 11.3 | 3,294 |
| Phase 5 | 901 | 26.1 | 616 | 17.8 | 919 | 26.6 | 624 | 18.1 | - | - | 392 | 11.4 | 3,452 |
| Phase 6 | 960 | 26.7 | 583 | 16.2 | 983 | 27.3 | 667 | 18.5 | - | - | 404 | 11.2 | 3,597 |
| Phase 7 | 962 | 26.1 | 644 | 17.5 | 1,038 | 28.1 | 684 | 18.5 | - | - | 360 | 9.8 | 3,688 |
| Phase 8 | 922 | 25.0 | 650 | 17.6 | 958 | 26.0 | 628 | 17.0 | - | - | 533 | 14.4 | 3,691 |
| Phase 9 | 995 | 25.6 | 676 | 17.4 | 1,071 | 27.6 | 687 | 17.7 | - | - | 455 | 11.7 | 3,884 |
| Phase 10 | 1,129 | 28.4 | 692 | 17.4 | 1,072 | 27.0 | 762 | 19.2 | 10 | 0.3 | 312 | 7.8 | 3,977 |
| Phase 11 | 1,033 | 26.4 | 730 | 18.7 | 1,079 | 27.6 | 723 | 18.5 | 25 | 0.6 | 320 | 8.2 | 3,910 |
| Total | 11,362 | 29.0 | 6,489 | 16.6 | 10,443 | 26.7 | 6,436 | 16.4 | 35 | 0.1 | 4,404 | 11.2 | 39,169 |
